# MULTISYSTEM INFLAMMATORY SYNDROME IN CHILDREN COURSE AMID COVID-19 PANDEMIC IN THE REBUBLIC OF BELARUS

**DOI:** 10.1101/2022.10.14.22280869

**Authors:** Oksana Romanova, Natalia Kolomiets, Dzianis Savitski, Ala Dashkevich, Olga Krasko, Galina Batyan, Anna Kluchareva, Marina Sokolova, Ekaterina Sergienko, Oksana Hanenko, Lydia Matush, Uladislava Senkevich

## Abstract

**Resume:** In first two years of COVID-19 pandemic, children usually had a mild or asymptomatic form of the disease. However, in rare cases, after suffering COVID-19, children had clinical manifestations similar to incomplete Kawasaki Disease (CD) or toxic shock syndrome. This condition is known as multisystem inflammatory syndrome in children (MIS-C).

**The purpose:** of this research was to study clinical and laboratory features and outcomes of multisystem inflammatory syndrome in children who were hospitalized during COVID-19 pandemic.

**Materials and methods:** In 19 months (May 2020 – December 2021) 63 patients with a diagnosis of “Multisystem inflammatory syndrome in children” (MIS-C) associated with COVID-19 were observed in the departments of Anesthesiology and Intensive Care of the Healthcare Institution “City Children’s Infectious Clinical Hospital” in Minsk, Republic of Belarus. MIS-C was diagnosed on criteria of CDC/WHO, 2020. All calculations were carried out in the statistical package R, version 4.1. The results of the analysis were considered statistically significant with p<0,05.

**The results of the study:** All of 63 children with MIS-C didn’t have an acute coronavirus infection. Therefore, it was impossible to determine which strain of SARS-CoV-2 patient exactly had. However, we formed 3 groups of patients based on circulation of the dominant strain of SARS-CoV-2 in Belarus at different times. The 1st group included 40 patients (63,5%) received treatment from 05.25.2020 to 02.21.2021 (“wuhan strains”); the 2nd group – 9 children (14,3%) from 02.23.2021 to 06.13.2021 (“alpha”); the 3rd group – 14 children (22,2%) from 07.01.2021 to 11.19.2021 (“delta”). 47 (74,6%) patients had complete and incomplete Kawasaki Disease phenotype of MIS-C. Nonspecific phenotype was observed in 16 (25,4%) children. It manifested as signs of shock. The mean age didn’t differ in study groups. It was 7±2,5; 9,4±4,2; 7,9±5 years respectively. All children had hyperthermic syndrome. Fever reached febrile numbers 3-4 times a day. Average fever duration was 3,2 [1–15] days. The course of MIS-C in children also didn’t depend on the circulating strain of the virus. For instance, gastrointestinal dysfunction was observed in all three groups with equal frequency (73%, 78% and 57%, respectively). The only a statistically significant increase was in the number of children with cheilitis. In the 2nd group 8 children (89%) and the 3rd group 13 children (93%) had cheilitis, respectively, p=0,002. Neurological disorders such as headache, hyperesthesia, hallucinations, photophobia were more often observed in the 1st group of children - 19 (48%) cases and less frequently in the 2nd and 3rd group (in 11% and 14% of cases), p=0,022. Pathological blood flow regurgitation was the most common disorder (68-71%). Several biochemical markers of inflammation levels, such as C-reactive protein (CRP) and procalcitonin (PCT), were high. CRP levels were 162 mg/l [130; 245]; 130 mg/l [90; 160]; 130 mg/l [106; 149] in 3 study groups, respectively. In children of the 1st group CRP level was significantly higher, p=0.052. PCT level was higher in patients of the 3rd group (4.2 ng/ml [2,4; 8,8]; 3.9 ng/ml [3,2; 11,9]; 8.7 ng/ml [3,4; 14,1], p=0.625).

**Conclusion:** As a result of the research there wasn’t found notable connection between clinical or laboratory features of MIS-C and the dominant circulating strain of SARS-CoV-2 in given time periods. During the circulation of “alpha” and “delta” strains, the only significant differences were decrease of the number of patients with neurological disorders and increase in the frequency of cheilitis, p=0,002. The remaining indicators of organ dysfunction were similar in three groups of children. There was 1 (1,6%) fatal outcome in our study.

## Introduction

At the end of 2019 a new coronavirus was identified. The spread of which quickly reached pandemic proportions [1]. At the beginning of the events, the children were not intensively involved in the epidemic process, usually the infection was mild or asymptomatic. But already in April 2020, reports from the United Kingdom recorded a clinic of a disease in children similar to incomplete Kawasaki Disease (KD) or toxic shock syndrome [2, 3]. Since then, there have been reports of the same clinical symptoms in children from other parts of the world [4-11]. This condition has been called MIS-C. It is also mentioned as pediatric multisystem inflammatory syndrome (PMI), pediatric inflammatory multisystem syndrome temporarily associated with SARS-CoV-2 (PIMS-TS), pediatric hyperinflammatory syndrome or pediatric hyperinflammatory shock. MIS-C is late immunopathological complication of COVID-19 in children and is characterized by fever, systemic vasculitis of likely autoimmune etiology with small and medium-sized vessels damage, with mandatory multisystem involvement of cardiovascular system, gastrointestinal tract, nervous system, kidneys, liver, eyes and skin. The incidence of MISC remains uncertain, although it is considered a relatively rare complication of COVID-19. It is observed in less than 1% of children with confirmed SARS-CoV-2 infection. According to Dallan C. et.al. [12, 13] in New York State, United States of America, the incidence of COVID-19 with laboratory confirmation of SARS-CoV-2 infection in people under the age of 21 is 322 per 100 000, and the incidence of MIS-C is 2 cases per 100 000 children.

The aim of the research is to study epidemiology, clinical and laboratory characteristics of MIS-C in patients, who were hospitalized in the period 2020-2021 in Belarus.

## Materials and methods

In 19 months (May 2020 – December 2021) 63 patients with a diagnosis of “Multisystem inflammatory syndrome in children” (MIS-C) associated with COVID-19 were observed in the departments of Anesthesiology and Intensive Care of the Healthcare Institution “City Children’s Infectious Clinical Hospital” in Minsk, Republic of Belarus. MIS-C was diagnosed on criteria of CDC/WHO [7, 11].

Parents of 24 (38%) children noted confirmed case of acute COVID-19 infection about a month before the development of MIS-C symptoms. The remaining 39 children sustained asymptomatic form of COVID-19 infection. All of the studied children had no significant comorbidity. Complete or incomplete CD and toxic shock syndrome (TSS) had been excluded in all hospitalized patients. Several laboratory tests had been performed: a general blood test with the calculation of platelet levels and formulas, erythrocyte sedimentation rate (ESR); biochemical study with determination of the level of C-reactive protein (CRP), urea, creatinine, liver functional tests (ALAT, ASAT); lactate dehydrogenase (LDH), ferritin, creatinine phosphokinase (CPK and CPK MB), protein, albumin, procalcitonin (PCC); coagulogram (prothrombin time, international normalized ratio, activated partial thromboplastin time); brain natriuretic peptide (BNP), NT-pro-BNP, N-terminal pro-BNP; chest radiography (CK); computer tomography (CT); electrocardiogram (ECG).

Virological examination for COVID-19 included determination of SARS-CoV-2 RNA in polymerase chain reaction (PCR); determination of specific IgM IgG to SARS-CoV-2 qualitatively and quantitatively in enzyme immunoassay (ELISA). To exclude Epstein-Barr virus (EBV); Cytomegalovirus (CMV), Parvovirus B19, Enterovirus infection and respiratory viruses PCR and ELISA methods were also used. Testing for other pathogenic microorganisms was carried out by generally accepted bacteriological methods. Biological material from the upper respiratory tract, blood, blood serum, feces had been used for immunological tests.

### Statistical methods of result processing

At first, the analysis of compliance of the kind of distribution of quantitative indicators with the law of normal distribution was carried out. It was performed using the Shapiro-Wilk criterion. Based on the results of the preliminary analysis, nonparametric methods of descriptive statistics were used in the calculations.

Quantitative indicators of the study are represented by median and quartiles in the form of median [Q25; Q75]. Comparison of quantitative indicators between three groups was carried out according to the Kruskal-Wallis criterion.

Qualitative indicators are represented by frequencies and percentages in the group. The chi-square criterion was used in the study of conjugacy tables. If chi-square criterion assumption was violated, the exact Fisher criterion had been used.

All calculations were carried out in the statistical package R, version 4.1. The results of the analysis were considered statistically significant with p<0,05 [14].

## Results and discussion

The first case of COVID-19 in the Republic of Belarus was registered on February 28, 2020 in an adult. According to epidemiological and molecular-genetic data, it was classified as imported from Iran.

The first sick child was identified 10 days after the registration of the first COVID-19 case. The infection occurred at the place of residence. It was a result of the implementation of the second generation of the infection spread cluster. From this point to the present, the epidemic process continues. It is characterized by a wave-like course and constant genetic variations switch-over of SARS-CoV-2. We evaluated events from February 28, 2020 to December 29, 2021, when the beginning of the circulation of “omicron” SARS-CoV-2 was officially announced for the first time [15].

According to the results of molecular genetic analysis, during the first year (February 2020 - February 2021), mainly four strains of WV SARS-CoV-2 (“wuhan strains”) circulated in Belarus. Then they were displaced by strain B.1.1.7 (“alpha”) SARS-CoV-2, which circulated until July 2021. The appearance of the next strain B.1.617.2 (“delta”) SARS-CoV-2 was accompanied by a high incidence and an increase in the number of hospitalizations, including hospitalization among children. [15]. “Delta” had been circulated almost till the end of December 2021. It is worth noting that vaccination of the adult population has been organized in the country since December 2020. Immunization of children 12-17 years old, though, has been started only since December 2021.

After the first case of MIS-C, children were admitted to treatment in a wavelike pattern, this dynamic is shown in Figure 1.

**Figure 1.**
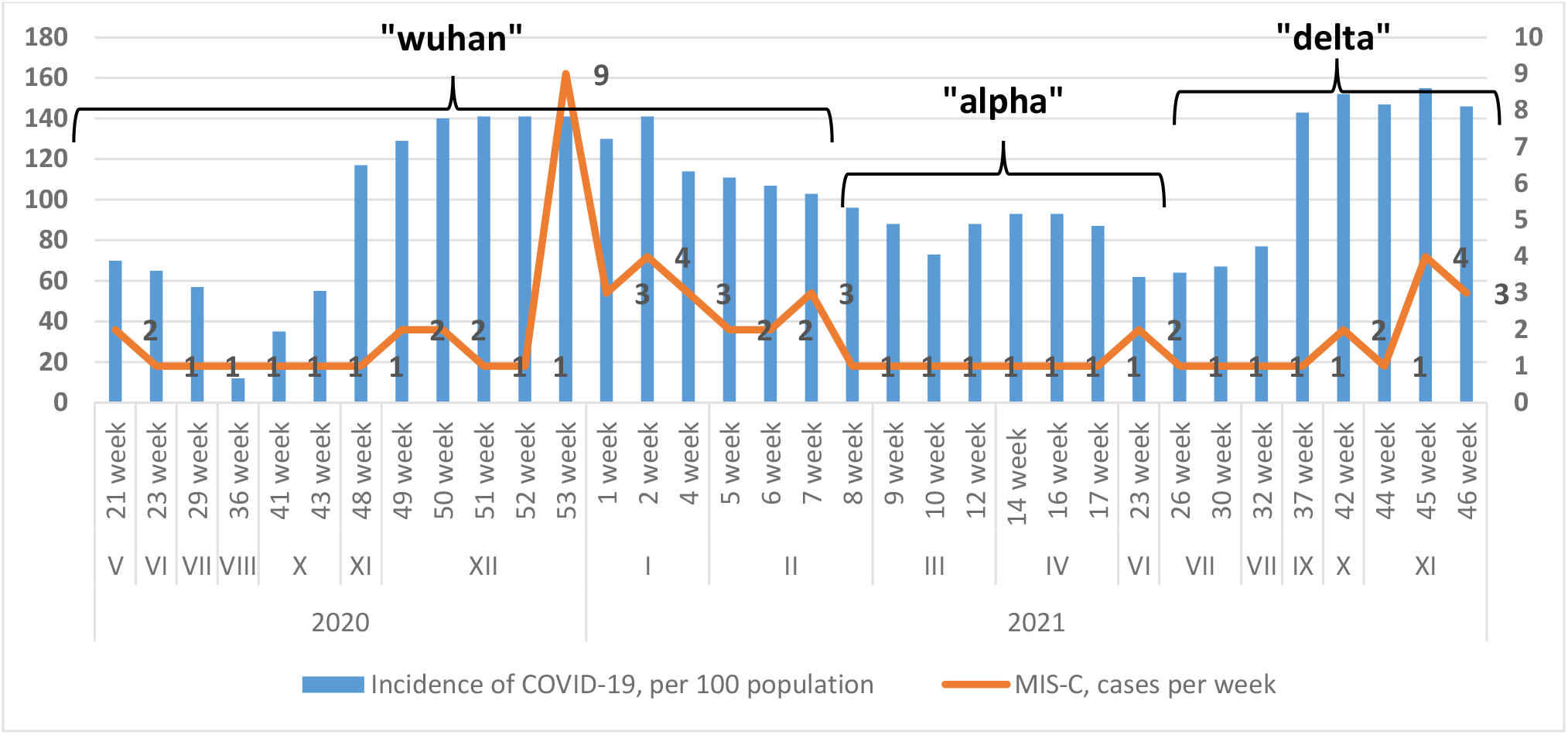
Dynamics of hospitalization of patients with MIS-C during 2020-2021

Naturally genetic mutations of viruses are not a new phenomenon and may go unnoticed. Nevertheless, sometimes virus can acquire new properties as a result of mutations, which change its characteristics. For instance, rate of spread, presence/absence of certain clinical signs, the severity of the disease, the effectiveness of certain drugs, vaccines might be changed.

All of 63 children with MIS-C didn’t have an acute coronavirus infection. Therefore, it was impossible to determine which strain of SARS-CoV-2 patient exactly had. Focusing on the periods of circulation of dominant coronaviruses, 3 groups of patients were formed. The 1st group included 40 patients (63,5%) received treatment from 05.25.2020 to 02.21.2021 (“wuhan strains”); the 2nd group – 9 children (14,3%) from 02.23.2021 to 06.13.2021 (“alpha”); the 3rd group – 14 children (22,2%) from 07.01.2021 to 11.19.2021 (“delta”).

In most studies, there was a gap of several weeks between a peak of acute COVID-19 in communities and an increase in MIS-C cases [4, 6, 10]. In our study, the first cases of COVID-19 in children requiring hospitalization began to be registered in April 2020. Against this background, the first child with MIS-C was admitted for treatment on May 19, 2021, that is, 82 days after the outbreak of the epidemic in the country, which is quite consistent with the literature data. For example, in London, the peak of COVID-19 cases occurred in the first to second weeks of April, while the spike in MIS-C cases occurred in the first to second week of May [16]. This three-to four-week break roughly coincides with the timing of acquired immunity and suggests that MIS-C may most likely represent a post-infectious complication rather than an acute COVID-19 infection in children. Although, according to most researchers, duration between acute infection and the appearance of the first symptoms of MIS-C is from 2 to 6 weeks.

There are reports about cases of MIS-C developing 6 weeks after acute SARS-CoV-2 infection. In a number of studies, the time between acute infection and the onset of MIS-C symptoms has not been determined, since most children probably suffered an asymptomatic form of infection. In later observations from the onset of the MIS-C pandemic, patients or their parents were more likely to know about their contact with COVID-19 and/or the date of positive testing. Apparently, increased epidemiological surveillance and broad public information about COVID-19 contributed to this [17]. In the Republic of Belarus, cases of COVID-19 were registered mainly among adult population, children accounted for an average of 10% in the structure of cases. Level of population immunity to the SARS-CoV-2 among belarusian population in the second year of the pandemic (May - September 2021) was quantified twice. In May 2021, level of seroprevalence among children aged 1-17 years was 39,2% [33,3; 41,6], in September 2021 – 25,4% [22,9; 28,1] [18].

According to gender structure, male children prevailed in all 3 groups: 28 (70%), 6 (66,7%) and 8 (57,1%). Their average age was 9 [2-12], 6 [1-14] and 7 [2-17] years, respectively. In 2nd and 3rd groups, the age of children was less than in 1st group, p= 0,235 (Table 1). All of the patients were Caucasians. Our findings are quite consistent with the results of a study conducted in the USA, which included 1,733 patients with MIS-C. There were 994 (57,6%) males, their average age was 9 [5-13] years [19].

**Table 1.** Anthropometric characteristics of patients with MIS-C

| Indicators | Total<br>n= 63 | 1<sup>st</sup> group<br>n=40 | 2<sup>nd</sup> group<br>n=9 | 3<sup>rd</sup> group<br>n=14 | p |
| --- | --- | --- | --- | --- | --- |
| Sex, n (%) |  |  |  |  |  |
| boys | 42 (67%) | 28 (70) | 6 (67) | 8 (57) | 0,68 |
| girls | 21 (33%) | 12 (30) | 3 (33) | 6 (43) |  |
| Age, month<br>median [Q 25; Q75] | 112<br>[72; 156] | 118<br>[85,5; 150] | 72<br>[52; 125,0] | 86<br>[61,2; 148,5] | 0,235 |

In available studies, the average age of children with MIS-C was 8-11 years in the range from 1 to 20 years, which is its distinguishing feature from KD. KD usually affects infants and young children, more often of Asian origin in East Asia [10, 19-22].

In our study, only 9 children (14%) had concomitant endocrine pathology (obesity). All other children were practically healthy, which corresponds to the literature data where in a number of observations most cases of MIS-C are diagnosed in older children and adolescents who were previously healthy, while severe acute COVID-19 develops in children with concomitant diseases such as endocrine pathology, malignant neoplasms, neurological, genetic diseases and others [19-23]. It is considered that MIS-C happens as a result of an abnormal immune response to a viral infection and has some clinical similarities with KD, macrophage activation syndrome (MAS) and cytokine release syndrome. Based on available studies, it is assumed that MIS-C appears to have an immunophenotype different from KD. However, the exact mechanisms of trigger an abnormal immune response by SARS-CoV-2 are still unknown and are being studied worldwide [24, 25].

Children with MIS-C were admitted on the 4nd [1-8] day after the appearance of the primary symptoms of the disease. Most of them were sent for hospitalization by primary care medical workers, emergency medical doctors or other hospitals. Only 10 (16%) applied independently.

50 of 63 patients (79,4%) had the MIS-C phenotype similar to complete and incomplete KD. Only 13 (20,6%) children had a nonspecific phenotype with signs of shock. At the same time, the incidence of cases of MIS-C, like complete and incomplete KD, increased as the dominant strain of SARS-CoV-2 changed. There were 29 (72,5%) in 1st group; 9 (100%) in 2nd group, and 12 (85,7%) in 3rd group of patients. At the same time there was a decrease in cases of phenotype with signs of shock – 11 (27,5%), 0 and 2 (14,3%) children, respectively.

In the light of experience on MIS-C, it becomes obvious that there is a wide range of interpretations of the severity of the disease. At the beginning of the pandemic, during the circulation of “Wuhan strains”, with a small number of cases, studies mainly reported the most severe course of MIS-C with a high incidence of shock (32-76%), myocardial damage (51-90%), arrhythmia (12%) and respiratory failure (28-52%). Recent publications report an increase of detection of milder forms of MIS-C, shock. Left ventricular dysfunction (LV) and respiratory failure also began occur less frequently [6, 13].

The clinical symptoms of MIS-C that we observed in our patients are presented in Table 2. As can be seen from the presented data, among the clinical symptoms of the disease, all children had hyperthermic syndrome with temperature rises to febrile figures 3-4 times a day and the duration of fever averaged 3,2 [1-15] days. 53 children (84%) were admitted to the hospital with a febrile temperature, the rest of the children had a subfebrile temperature. According to a number of studies, fever lasts from 3 to 5 days in most patients [6, 13].

**Table 2.**
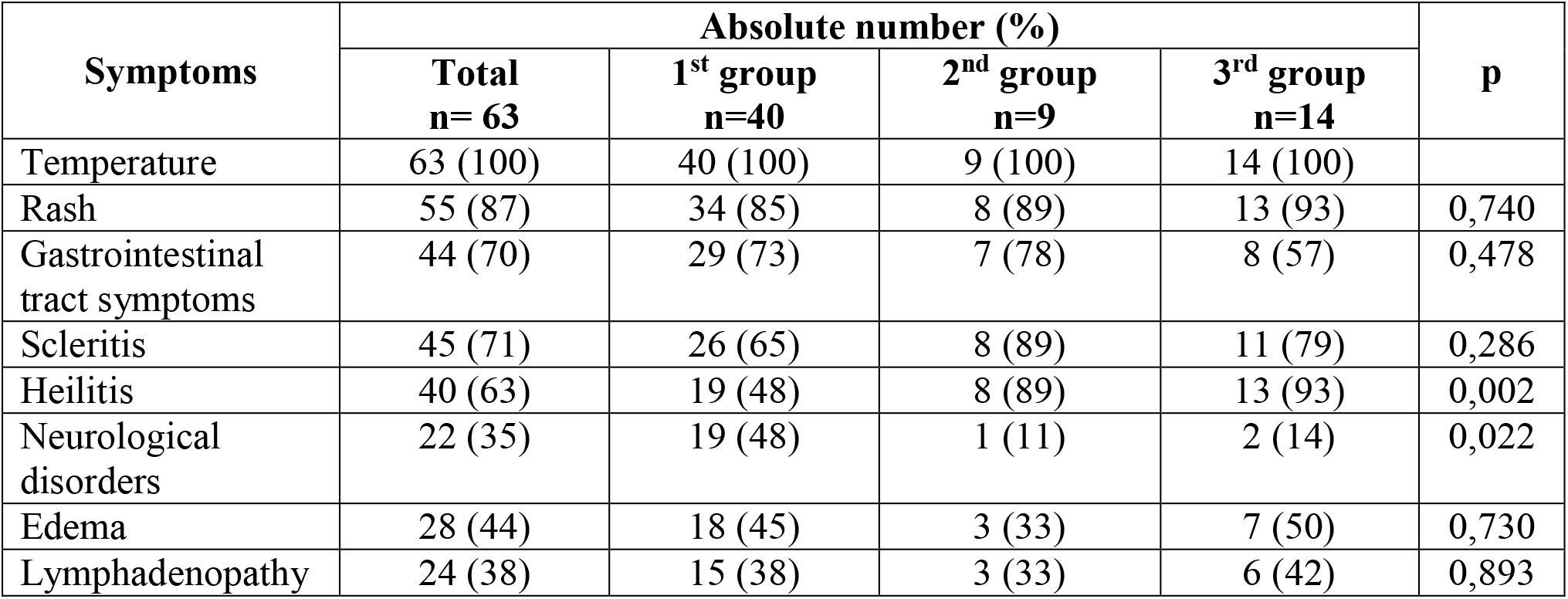
Clinical symptoms of MIS-C in children

In the research involving 186 patients, it was described that 10% of patients had fever for 3 days, 12% for 4 days and 78% for 5 or more days [6]. In an American study involving 1733 patients with MIS-C, the median duration of fever was 5 [4-7] days [19].

On the 2nd-4th days of the disease, the appearance of a large- or small-spotted rash prone to fusion, without typical localization sites, occurring with the same frequency (85%, 89% and 93%, p=0,740), regardless of circulating strains of SARS-CoV-2 was observed. Rarely there was a small-point, hemorrhagic rash. On the background of therapy, the rash passed on average by 3 [1-5] days. According to other researchers, rash in children with MIS-C is observed in 45-76% of patients [4, 9, 26, 27]. Mucosal lesions (scleritis, cheilitis) we observed diseases on 2-3 days, which on average persisted for up to 3 [1-6] days.

We noted a statistically significant (p=0,002) increase in the frequency of manifestations of cheilitis (red and swollen lips, raspberry tongue) in children, depending on the isolated COVID-19 strains, and an increase in the number of children with cheilitis during the “alpha” and “delta” circulation compared to the “wuhan strains” of coronavirus – 19 (48%), 8 (89%), 13 (93%) accordingly. For children with scleritis, it was also higher in children in 2nd group (8 (89%) and 3rd group – 11 (79%), in contrast to 1st group – 26 (65%) (p=0,286). Similar data were published by other researchers who observed scleritis in 30-81% of patients with MIS-C, and cheilitis in 27-76% of children [4, 20, 26].

According to literary sources, sometimes children have lesions of the mucous membrane of the genitals, leading to dysuric disorders against this background. In these cases, a urinary catheter is required to relieve urinary retention syndrome. Due to swelling of the urinary tract mucosa, we performed the placement of a urinary catheter in 2 children (3%). In this study, hourly diuresis during the first knocks of the stay was preserved or reduced to oliguria, however, normalization was noted against the background of therapy by the second day.

Gastrointestinal tract dysfunction was manifested by pain syndrome, diarrhea, vomiting, decreased appetite and was observed in our study more often in children during the circulation of “wuhan strains” – 29 (73%) and “alpha” – 7 (78%), was less often recorded in children who fell ill during the circulation “delta” – in 8 (57%) cases (p=0,478). The duration of the observed symptoms in our study averaged 2 days [1-6 days]. In 2nd group, one patient with MIS-C abdominal pain was simulated appendicitis, which required transfer to the surgical department for laparotomy. According to a number of studies, gastrointestinal symptoms such as abdominal pain, vomiting, diarrhea are especially common and clearly manifest and mimic appendicitis. It is also described that some children developed terminal ileitis during abdominal imaging and/or colitis during colonoscopy [28]. In the study by Belay E.D. et al. [19], which included 1733 patients with MIS-C, in addition to fever, the most common signs and symptoms were abdominal pain −1153 (66,5%), vomiting −1114 (64,3%), rash – 963 (55,6%), diarrhea – 931 (53,7%) and conjunctival hyperemia – 929 (53,6%). In this study, interaction was established between the presence of symptoms and the age of the observed. Thus, patients aged 0 to 4 years had the lowest proportion of lesions of more than 6 organ systems, as well as gastrointestinal symptoms, hypotension, shock, myocarditis, cardiac dysfunction, so they were less likely to be hospitalized in the intensive care unit.

Neurological disorders such as headache, hyperesthesia, hallucinations, photophobia were more often observed in our study in 19 (48%) children from 1nd group (p=0,022) and less often in 2nd and 3rd groups – 1 (11%) and 2 (14%) cases, accordingly. According to a number of studies, neurocognitive symptoms are common, observed in 29-58% of patients and include headache, lethargy, confusion or irritability. However, a small number of patients have severe neurological manifestations, which include encephalopathy, attack, coma, stroke, meningoencephalitis, muscle weakness and signs of damage to the brain stem and/or cerebellum [16, 24, 36]. In a study involving 616 patients with MIS-C, 20% had neurological damage. Life-threatening neurological conditions occurred only in 20 (3%) patients, and included severe encephalopathy (n=8), demyelination of the central nervous system (n=6), stroke (n=3), acute lightning brain edema (n=2) and Guillain-Barre syndrome (n=1) [6, 29, 30].

Lymphadenopathy and edematous syndrome in our study were observed with the same frequency (38%, 33% and 42%, p=0,893) and (45%, 33% and 50%, p=0,730), respectively, regardless of the circulation of the dominant strains of SARS-COV-2. We noticed that according to the literature data, lymphadenopathy and edematous syndrome were observed significantly less frequently - 6-16% and 9-16% of children with MIS-C, respectively [4, 27].

Heart damage, according to a number of studies, is common and is a characteristic sign of MIS-C. The mechanisms of myocardial damage in MIS-C are not yet well describe. Possible causes include damage as a result of systemic inflammation, acute viral myocarditis, hypoxia, stress cardiomyopathy and, in rare cases, ischemia caused by damage to the coronary artery (CA). Cardiac dysfunction (CV) in some patients may be the result of a combination of these mechanisms. Given the variability of clinical manifestations, it is likely that different mechanisms are responsible for the defeat of the cardiovascular system in different patients. There are limited research data characterizing the histopathology of the heart in MIS-C. In the report of one study on a fatal case of MIS-C, signs of myocarditis, pericarditis and endocarditis characterized by infiltration of inflammatory cells were noted in the autopsy results [6, 31-33]. In addition, the SARS-CoV-2 virus was detected in the heart tissue using electron microscopy and PCR. However, some clinical signs in this patient were uncharacteristic of MIS-C and, first of all, there was severe lung damage, and it is quite possible that the autopsy results more closely reflect the severe acute form of COVID-19, and not MIS-C. In some large series of studies, approximately 30 to 40% of children had suppressed LV (left ventricular) function and 8 to 24% had KA abnormalities [4, 13, 19, 25]. These studies included patients with both severe MIS-C and milder cases. In studies that concerned only seriously ill patients, significantly higher rates of suppressed LV function (approximately 50 to 60%) and KA anomalies (approximately 20 to 50%) were shown. As discussed in the literature, heart failure is a key feature that helps distinguish MIS-C from severe acute COVID-19 [24, 32].

When analyzing the heart ultrasound indicators of our patients, we noticed that pathological regurgitation of blood flow is the most frequent disorders. For example, the mitral valve (MV) was recorded in 43 (68%) of the observed, but it was more common in patients, during the circulation of “wuhan strains” – 31 (78%). Regurgitation of blood flow on the pulmonary valve (PV) and tricuspid valve (TV) was also observed in the majority of patients – 44 (69,8%) and 47 (74,6%) children, respectively, in the case of regurgitation on aortal valve (AV) in only 4 patients of group 1. LV dilatation was detected equally frequently regardless of the circulating strain and totaled 32 (50,8%) observations. Dilation of the left and right coronary arteries (CA) was observed in 26 (41,3%) children, but during the “delta” circulation the proportion of violations was 71%. Effusion into the right and left pleural cavity was observed in 24 (38%) and 28 (44%) cases, respectively. In contrast to the above data on these indicators, statistically significant differences were noted in the 2nd and 3rd groups p=0,033 (Table 3).

**Table 3.**
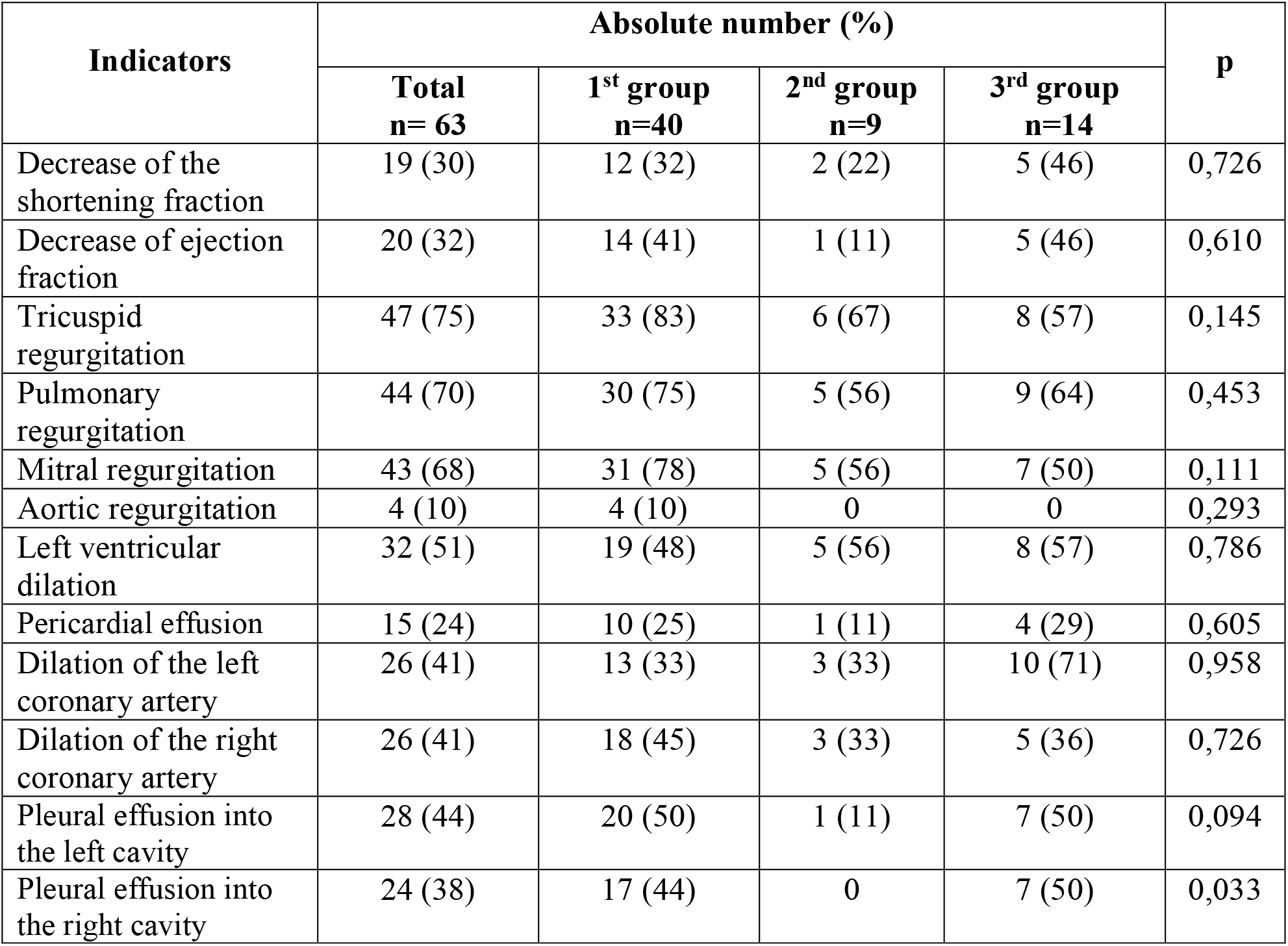
Cardiac manifestations of MIS-C in patients

| Indicators | Absolute number (%) |  |  |  | p |
| --- | --- | --- | --- | --- | --- |
|  | Total<br>n= 63 | 1<sup>st</sup> group<br>n=40 | 2<sup>nd</sup> group<br>n=9 | 3<sup>rd</sup> group<br>n=14 |  |
| Decrease of the shortening fraction | 19 (30) | 12 (32) | 2 (22) | 5 (46) | 0,726 |
| Decrease of ejection fraction | 20 (32) | 14 (41) | 1 (11) | 5 (46) | 0,610 |
| Tricuspid regurgitation | 47 (75) | 33 (83) | 6 (67) | 8 (57) | 0,145 |
| Pulmonary regurgitation | 44 (70) | 30 (75) | 5 (56) | 9 (64) | 0,453 |
| Mitral regurgitation | 43 (68) | 31 (78) | 5 (56) | 7 (50) | 0,111 |
| Aortic regurgitation | 4 (10) | 4 (10) | 0 | 0 | 0,293 |
| Left ventricular dilation | 32 (51) | 19 (48) | 5 (56) | 8 (57) | 0,786 |
| Pericardial effusion | 15 (24) | 10 (25) | 1 (11) | 4 (29) | 0,605 |
| Dilation of the left coronary artery | 26 (41) | 13 (33) | 3 (33) | 10 (71) | 0,958 |
| Dilation of the right coronary artery | 26 (41) | 18 (45) | 3 (33) | 5 (36) | 0,726 |
| Pleural effusion into the left cavity | 28 (44) | 20 (50) | 1 (11) | 7 (50) | 0,094 |
| Pleural effusion into the right cavity | 24 (38) | 17 (44) | 0 | 7 (50) | 0,033 |

In an international study involving 503 patients with MIS-C, during echocardiography, 34% had a decrease in LV ejection fraction, 13% had KA aneurysms [19]. Among patients with suppressed LV function, 55% had mild LV depression, moderate depression in 23% and severe in 22% of children. The majority of KA aneurysms (93%) were of mild severity, 7% were of moderate severity, and no large or giant KA aneurysms were detected in any case [20]. In 91% of patients, LV function returned to normal within 30 days, and in almost all patients with available 90-day follow-up data, LV function became normal. The outcomes of CA aneurysms were equally favorable, regressing to normal (Z-score <2,5) in more than three quarters of affected patients within 30 days and in all patients with available 90-day follow-up data [20]. In our study, against the background of ongoing therapy in all discharged children, these changes from the cardiovascular system were reversible.

The ECG findings presented in Table 4 demonstrate myocardial repolarization disorders in the groups of 45%, 33%, 50%; bradyarrhythmia of 15%, 22%, 29%; AV blockade of the 1st degree of 10%, 11%, 7% and migration of the pacemaker of 5%, 33%, 7%, respectively.

**Table 4.** ECG disorders of MIS-C patients

| Indicators | Absolute number (%) |  |  |  |
| --- | --- | --- | --- | --- |
|  | Total<br>n= 63 | 1<sup>st</sup> group<br>n=40 | 2<sup>nd</sup> group<br>n=9 | 3<sup>rd</sup> group<br>n=14 |
| Repolarization disorders | 28 (44) | 18 (45) | 3 (33) | 7 (50) |
| Bradyarrhythmia | 12 (19) | 6 (15) | 2 (22) | 4 (29) |
| AV block (1st degree) | 6 (9,5) | 4 (10) | 1 (11) | 1 (7) |
| Migration of the pacemaker | 6 (9,5) | 2 (5) | 3 (33) | 1 (7) |

Upon admission to the hospital, almost all children had a compensated acid-base state, which averaged 7,39 [7,1-7,52], with glycemic indices averaging 6,1 [4,1–13,2] mmol/L and lactate - on average 1,9 [1-10] mmol/L. According to the electrolyte composition of the blood, hyponatremia was registered in 18 children (29%), the others indicators were within the normal range.

Changes in the clinical blood analysis are presented in Table 5. We noted that leukocytosis was more often observed in children in the 1st (40%) and 3rd groups (43%), less often in the 2nd (22%), p=0,224, leukopenia was more often observed in the third group – in 21% of patients, neutrophilia (65%, 67% and 64%, respectively, p=0.951) and lymphocytopenia (68%, 67% and 64%, respectively, p=0,976) were observed equally frequently regardless of the current COVID-19 pandemic. The maximum indicators of the accelerated erythrocyte sedimentation rate (ESR) were reached by the 7-9 day of hospital stay. Anemia was increasing by the 2-3 day of hospital stay. Thrombocytopenia (less than 150*10^9^/l) at admission was observed more often in the 1st and 3rd groups (35% and 36%, respectively) and less often in the 2nd – 11%, p=0,597. We found no statistically significant differences in the hemogram depending on the current COVID-19 pandemic.

**Table 5.**
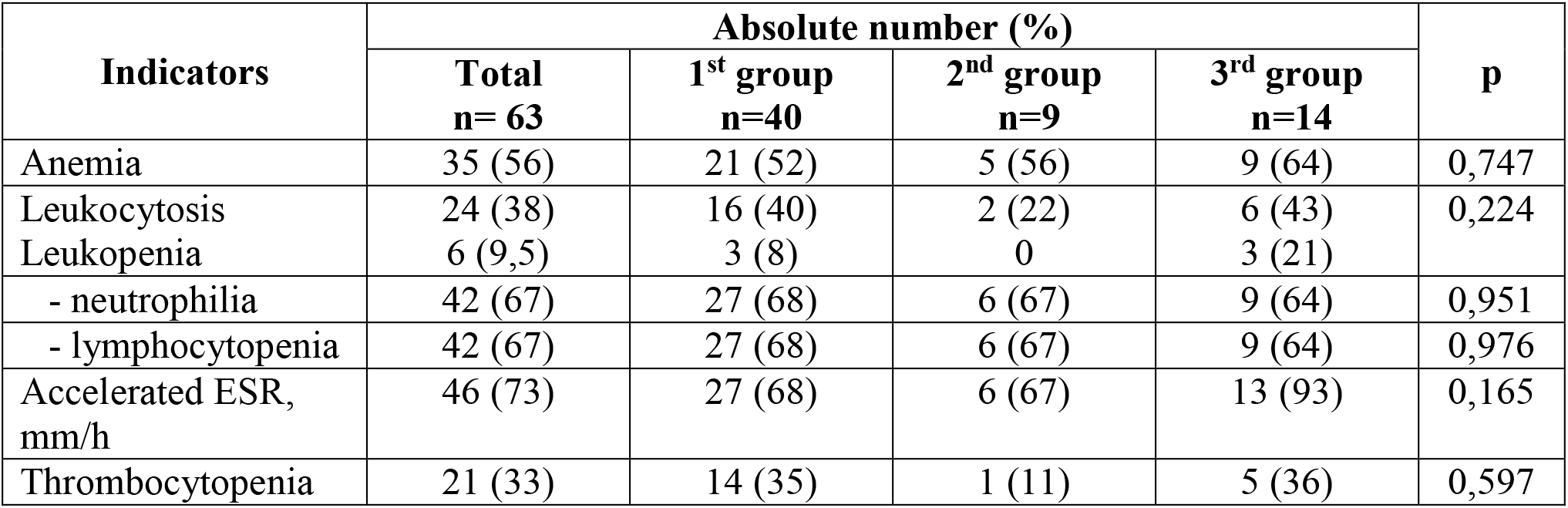
Hemogram indicators of MIS-C patients

| Indicators | Absolute number (%) |  |  |  | p |
| --- | --- | --- | --- | --- | --- |
|  | Total<br>n= 63 | 1<sup>st</sup> group<br>n=40 | 2<sup>nd</sup> group<br>n=9 | 3<sup>rd</sup> group<br>n=14 |  |
| Anemia | 35 (56) | 21 (52) | 5 (56) | 9 (64) | 0,747 |
| Leukocytosis | 24 (38) | 16 (40) | 2 (22) | 6 (43) | 0,224 |
| Leukopenia | 6 (9,5) | 3 (8) | 0 | 3 (21) |  |
| - neutrophilia | 42 (67) | 27 (68) | 6 (67) | 9 (64) | 0,951 |
| - lymphocytopenia | 42 (67) | 27 (68) | 6 (67) | 9 (64) | 0,976 |
| Accelerated ESR, mm/h | 46 (73) | 27 (68) | 6 (67) | 13 (93) | 0,165 |
| Thrombocytopenia | 21 (33) | 14 (35) | 1 (11) | 5 (36) | 0,597 |

**Table 6.** Indicators of biochemical blood analysis of MIS-C patients

| Indicators | median [Q 25; Q75] / n (%) |  |  | P |
| --- | --- | --- | --- | --- |
|  | 1<sup>st</sup> group<br>n=40 | 2<sup>nd</sup> group<br>n=9 | 3<sup>rd</sup> group<br>n=14 |  |
| Albumin, mmol/l<br>- ↓ level | 39 [33; 41]<br>9 (23) | 39 [36; 40]<br>- | 36 [32; 40]<br>2 (14) | 0,461 |
| Urea, mmol/l<br>- ↑ level | 3,8 [3,4; 5,0]<br>3 (7,5) | 3,1 [2,8; 3,8]<br>- | 4,8 [3,8; 6,1]<br>1 (7) | 0,054 |
| Creatinine, mcmol/l<br>- ↑ level | 68 [49; 77]<br>2 (5) | 54 [44; 67]<br>- | 60 [56; 80]<br>1 (7) | 0,384 |
| ALT, E/l<br>- ↑ level | 36 [24; 52]<br>10 (25) | 27 [26; 67]<br>1 (11) | 38 [21; 56]<br>6 (43) | 0,989 |
| AST, E/l<br>- ↑ level | 38 [28; 57]<br>11 (28) | 36 [27; 56]<br>2 (22) | 36 [28; 50]<br>5 (36) | 0,930 |
| LDH, E/l<br>- ↑ level | 508 [380; 594]<br>18 (45) | 534 [463; 614]<br>5 (56) | 474 [408; 551]<br>8 (57) | 0,688 |
| CPK, E/l<br>- ↑ level | 64 [32; 166]<br>7 (16) | 47 [30; 112]<br>1 (11) | 95 [70; 186]<br>3 (42) | 0,361 |
| CPK MB, E/l<br>- ↑ level | 19 [16; 25]<br>9 (18) | 24 [17; 37]<br>3 (33) | 24 [20; 29]<br>6 (43) | 0,074 |
| CRP, mg/l<br>- ↑ level | 162 [130; 245]<br>40 (100) | 130 [90; 160]<br>9 (100) | 130 [106; 149]<br>14 (100) | 0,052 |
| PCT, ng/ml<br>- ↑ level | 4,2 [2,4; 8,8]<br>40 (100) | 3,9 [3,2; 11,9]<br>9 (100) | 8,7 [3,4; 14,1]<br>14 (100) | 0,625 |
| D-Dimer, ng/ml<br>- ↑ level | 1010 [664; 1972]<br>38 (95) | 919 [777; 1702]<br>9 (100) | 1100 [1052; 2230]<br>14 (100) | 0,834 |
| Fibrinogen, g/l<br>- ↑ level | 9,6 [7,5; 11,8]<br>40 (100) | 9,4 [8,6; 13,6]<br>8 (89) | 6,1 [5,0; 10,6]<br>1 (93) | 0,131 |

According to the literature sources, lymphopenia occurs from 80-95%, neutrophilia – 68-90%, anemia of mild degree – 70% and thrombocytopenia – 31-80%, which is consistent with the data of our study [4, 6, 13, 23, 35-37].

In the biochemical analysis of blood, we noted an increase in the levels of inflammatory markers: CRP (162 mg/l [130; 245]; 130 mg/l [90; 160]; 130 mg/l [106; 149], p=0,052) was recorded higher in children in the 1st group. PCT was higher in patients in the 3rd group (4.2 ng/ml [2,4; 8,8]; 3,9 ng/ml [3,2; 11,9]; 8,7 ng/ml [3,4; 14,1], p=0,625). There were no differences between the groups in other indicators of biochemical analysis.

According to literature data, laboratory markers of inflammation correlate with the severity of the disease [35-37]. In a number of research, increased markers of inflammation were observed, such as CRP – from 90 to 100%, accelerated ESR – from 75 to 80%, high levels of D-dimers – from 67 to 100%, fibrinogen – from 80 to 100%, ferritin – from 55 to 76%, procalcitonin – from 80 to 95% and interleukin-6 (IL-6) – from 80 to 100% [4, 16, 18, 21, 23, 31, 35]. In our research, children in all groups had high rates of CRP of 162 mg/l [130; 245]; 130 mg/l [90; 160]; 130 mg/l [106; 149], p=0,052), respectively, while patients of 1st group were distinguished. The marker PCT was higher in patients in the 3rd group (4,2 ng/ml [2,4; 8,8]; 3,9 ng/ml [3,2; 11,9]; 8,7 ng/ml [3,4; 14,1], p=0,625). Although other biochemical parameters had high values in relation to the reference ones, there were no differences in data between the groups.

According to chest radiography/computed tomography of the chest organs, 28 (44%) of children had lung changes in the form of interstitial changes or pneumonia. Of these, 5 (17%) of children had signs of respiratory insufficiency of the 2nd degree and needed oxygen therapy with an average duration of 3 [1-5] days. According to ultrasound of the pleural cavities, 25 (40%) children had an effusion in the right pleural cavity, 28 (44%) in the left pleural cavity. 4 (6%) children with MIS-C needed artificial ventilation of the lungs.

The fatal outcome in our study was observed in 1 (1,6%) adolescent patient in the 1st wave, who was admitted to the hospital with signs of shock, severe hypotension, cardiovascular and respiratory insufficiency and concomitant endocrine pathology. The autopsy revealed signs of congenital pathology on the part of the cardiovascular system. In a large American study involving 1733 patients, a fatal outcome was observed in 24 (1,4%) patients. Also in this study, an association with age was revealed, and the highest mortality was observed in patients aged 15 to 17 years and amounted to 2,6% [19].

## Conclusion

In our study, we observed 63 children with MIS-C ages 1 to 17 amid COVID-19 pandemic. The data of surveillance system shows that during 19 months of our observation (May 2020 – December 2021) on the territory of Belarus there were 3 changes of the dominant SARS-CoV-2 strain. They were “wuhan strains” (February 2020 - February 2021), “alpha” (February 2021 - July 2021), “delta” (July 2021 – December 2021). The first cases of MIS-C in children began to be registered approximately 2,7 months after the outbreak of the epidemic in the country and a month after the detection of severe COVID-19 in children, which is quite consistent with the data of other studies [16, 37, 38]. The largest number of patients – 40 (66,7%), were admitted to inpatient treatment during the circulation of the “wuhan strains” of SARS-CoV-2. Perhaps this was due to the long duration of this period and insufficient alertness of parents. Clinically predominant MIS-C phenotype was similar to complete and incomplete KD. It was diagnosed in 47 (74,6%) children. Nonspecific phenotype in the form of signs of shock was in 16 (25,4%). The average age in three groups did not differ and was 7±2,5; 9,4±4,2; 7,9±5 years, respectively. This is consistent with the literature data, where most cases of MIS-C occurred in older (≥5 years) and previously healthy children. We drew attention to the fact that only 9 (14%) of 63 children had concomitant endocrine pathology (obesity). All children had hyperthermic syndrome with rises in temperature to febrile figures 3-4 times a day and fever duration on average 3,2 [1–15] days. Clinical symptoms of MIS-C – rash, gastrointestinal dysfunction, scleritis, edematous syndrome, lymphadenopathy was observed in most children, but had no pronounced differences in the groups we formed. However, cheilitis developed in 40 (63%) of their 63 children was significantly more common (p=0,002) during the “alpha” and “delta” circulation periods”, 8 (89%) and 13 (93%) children, respectively. Disorders of the nervous system occurred less frequently in total – only in 22 (35%) children. 19 of them were observed during the circulation of “wuhan strains”, p=0,022. They manifested in the form of headaches, hyperesthesia, hallucinations, photophobia. The analysis of the heart ultrasound of the patients showed that pathological regurgitation of blood flow is the most frequent violation. The only statistically significant indicator was effusion into the right pleural cavity. We have seen it more often in the 3rd group, p=0,033. Increased levels of inflammatory markers, changes in the number of blood cells and other changes were noted in the blood tests. However, these changes were approximately equally often in all groups of patients. Despite the rather severe course of the disease, we had only 1 (1,6%) fatal outcome out of 63 children with MIS-C. It had happened at the initial stage of the study during the circulation of “wuhan strains”. The clinical course, diagnosis and treatment of MIS-C in children continue to be under scrutiny of specialists.

## Data Availability

All data produced in the present work are contained in the manuscript

